# Cerebral Small Vessel Disease and Cognitive Decline Following Transient Ischemic Attack: A Longitudinal Study

**DOI:** 10.1101/2025.03.05.25323459

**Authors:** Paula Roesen, Uchralt Temuulen, Ana Sofia Rios, Ramanan Ganeshan, Tim Bastian Braemswig, Ahmed Khalil, Kersten Villringer, Thomas Ihl, Huma Fatima Ali, Pimrapat Gebert, Ulrike Grittner, Michael Ahmadi, Matthias Endres, Heinrich J Audebert, Anna Kufner

**Affiliations:** Charité – Universitätsmedizin Berlin, corporate member of Freie Universität Berlin and Humboldt-Universität zu Berlin, Klinik für Neurologie mit Experimenteller Neurologie, 12203 Berlin, Germany; Charité – Universitätsmedizin Berlin, corporate member of Freie Universität Berlin and Humboldt-Universität zu Berlin, Center for Stroke Research Berlin (CSB), 12203 Berlin, Germany; Klinik und Hochschulambulanz für Neurologie, Campus Benjamin Franklin, Charité Universitätsmedizin Berlin, Berlin, Germany; Berlin Institute of Health, Berlin, Germany; Charité – Universitätsmedizin Berlin, corporate member of Freie Universität Berlin and Humboldt-Universität zu Berlin, Institute of Biometry and clinical Epidemiology, Charitéplatz 1, 10117 Berlin, Germany; Berlin Institute of Health at Charité – Universitätsmedizin Berlin, 10117 Berlin, Germany; German Center for Cardiovascular Research (Deutsches Zentrum für Herz Kreislauferkrankungen, DZHK), Partner Site Berlin, 10117 Berlin, Germany; German Center for Neurodegenerative Diseases (Deutsches Zentrum für Neurodegenerative Erkrankungen, DZNE), Partner Site Berlin, 10117 Berlin, Germany; German Center for Mental Health (DZPG), partner site Berlin, 10117 Berlin, Germany; Charité – Universitätsmedizin Berlin, corporate member of Freie Universität Berlin and Humboldt-Universität zu Berlin, Einstein Center for Neurosciences (ECN), 10117 Berlin, Germany

**Keywords:** Cerebrovascular risk factors, Cognitive impairment, Cerebral small vessel disease, Transient ischemic attack

## Abstract

**Background:** Cerebral small vessel disease (CSVD) is a common incidental finding on cerebral MRI in patients with transient ischemic attack (TIA) and stroke and has been linked to increased cerebrovascular risk and cognitive decline. This study aimed to investigate the prevalence of CSVD imaging biomarkers in TIA patients and evaluate their association with cognitive function over three years following the ischemic event.

**Methods:** A cohort of 246 TIA patients from the INSPiRE-TMS (ClinicalTrials.gov: NCT01586702) study were included. The CSVD-score – including white matter hyperintensities (WMH), lacunes, cerebral microbleeds (CMBs), and enlarged perivascular spaces (PVS) – was assessed on baseline MRI. Cognitive performance was assessed via the Montreal Cognitive Assessment (MoCA) at baseline and annual outpatient visits up to 3 years.

**Results:** CSVD was present in 58.5% of TIA patients. The most prevalent imaging biomarker was lacunes (36.6%), followed by PVS (28.1%), WMH (19.5%) and CMBs (17.9%). Cumulative CSVD-score (range 0-4) was an independently associated with cognitive decline up to 3 years (β = -0.53, 95% CI -0.97 – -0.09, p = 0.018), alongside advanced age (β = -0.08, 95% CI -0.13 – -0.03, p=0.001). CMB burden was the strongest predictive component of the CSVD-score (β = 0.42, 95% CI -0.63 – -0.21, p < 0.001). Specifically, CSVD-score had a significant negative effect on the *memory* domain of cognitive function with an adjusted β of - 0.18 (95% CI -0.32 – -0.04, p = 0.014).

**Conclusion:** Imaging biomarkers of CSVD are present in more than half of TIA patients and are an independent predictor of cognitive decline up to 3 years, with the strongest effect on the *memory* domain of cognitive function. Whether the presence of CMBs is the strongest predictive imaging biomarker of cognitive decline in TIA patients requires confirmation in further studies.

## Introduction

Cognitive decline affects one in five patients globally and represents a significant global health challenge; not only does it negatively affect patient quality of life, but it also poses a substantial socioeconomic burden, especially in an ageing global population.^1^ Although cognitive decline can have many etiologies - including neurodegenerative diseases, systemic conditions, and vascular pathologies - cerebrovascular disease is one of the leading causes of cognitive impairment in patients over the age of 75.^2,3^ Cerebrovascular disease not only exacerbates neurodegenerative processes but also independently leads to cognitive deficits by impairing cerebral perfusion and affecting neural networks.^4,5^

Cerebral small vessel disease (CSVD) is often an incidental finding on cerebral magnetic resonance imaging (MRI) in patients following an acute cerebrovascular event.^6^ Imaging biomarkers of CSVD include white matter hyperintensities (WMH), lacunes, cerebral microbleeds (CMBs), and enlarged perivascular spaces (PVS).^7^ Recent, large-scale studies including over 70,000 stroke patients have found that concomitant CSVD is a significant predictor of future cognitive decline.^8,9^ This trend has also been observed in smaller cohorts of transient ischemic attack (TIA) patients.^10^

Recent studies in mixed cohorts examining stroke and TIA patients reveal that up to 35% of patients experience cognitive decline within one year depending on the severity of the initial cerebrovascular event.^11,12^ Previous population-based studies have shown that CSVD is an independent predictor of cognitive decline, as well as overall long-term vascular risk and mortality.^8,9^ However, most studies include both ischemic stroke patients and TIA patients. Although such studies are valuable because ischemic stroke and TIA patients have similar risk profiles, it does now allow for the distinction between the effect of the acute ischemic lesion on cognitive decline and CSVD alone. To the best of our knowledge, there have been no studies as of yet that have investigated the effect of CSVD on long-term cognitive trajectory in a comprehensive cohort of TIA patients.

Therefore, we set out to investigate the prevalence of CSVD markers assessed on baseline MRI in a high-risk TIA population, and whether CSVD is associated with cognitive decline up to three years after the acute ischemic event. Furthermore, we investigated whether the severity of CSVD leads to differential decline across cognitive subdomains (i.e. *visuospatial/ executive abilities, naming, attention, abstraction, memory,* and *orientation*) following TIA.

## Materials and Methods

### Participants

This is a post-hoc analysis of a subset of data from the INSPiRE-TMS study (Intensified Secondary Prevention Intending a Reduction of Recurrent Events in TIA and Minor Stroke Patients; ClinicalTrials.gov: NCT01586702). Briefly, INSPiRE-TMS was an open-label, multicenter, randomized controlled trial that aimed to explore the effects of an intensified aftercare program in stroke and TIA patients. The main study inclusion criteria were patients with ischemic and hemorrhagic non-disabling stroke (modified Rankin Scale [mRS] score ≤2) or high-risk TIA (defined as ABCD2 score ≥ 3)^13^ within two weeks of study enrolment and the presence of at least one modifiable risk factor (i.e. arterial hypertension, diabetes, atrial fibrillation, or smoking). Patients were randomised either to an intensified support programme including up to eight outpatient visits over 2 years including motivational interviewing strategies or to conventional care alone. For a detailed description of the main trial inclusion and exclusion criteria, please refer to Ahmadi et al. 2020.^14^

For the current analysis, patients were included if they had (1) an available MRI performed as part of the clinical routine diagnostic workup during the acute hospital stay, (2) the clinical diagnosis of a TIA with (3) no diffusion restriction present on baseline MRI and (4) received assessment of cognitive status on at least one time-point within 3 years following the index event.

Of the 2098 patients enrolled in the INSPiRE-TMS trial, 797 were initially classified as a TIA based on initial clinical presentation and computed tomography (CT) imaging. Of those, 365 received an MRI and 121 of these showed acute diffusion restriction on the MRI and were subsequently diagnosed with ischemic stroke. Two patients were initially diagnosed with ischemic stroke at the time of study enrolment but were then diagnosed with TIA following further diagnostic work-up. For a detailed flow-chart of patients included/ excluded in the current study, please refer to **Figure 1**. In total, 246 patients were included in the final analysis. Out of 246 patients, 100 had Montreal Cognitive Assessments (MoCAs) up to 3 years follow-up with at least one value (98 patients with baseline MoCA, 88 with 1 year MoCAs, 79 with 2 year MoCAs and 64 with 3 year MoCAs).

**Figure 1:**
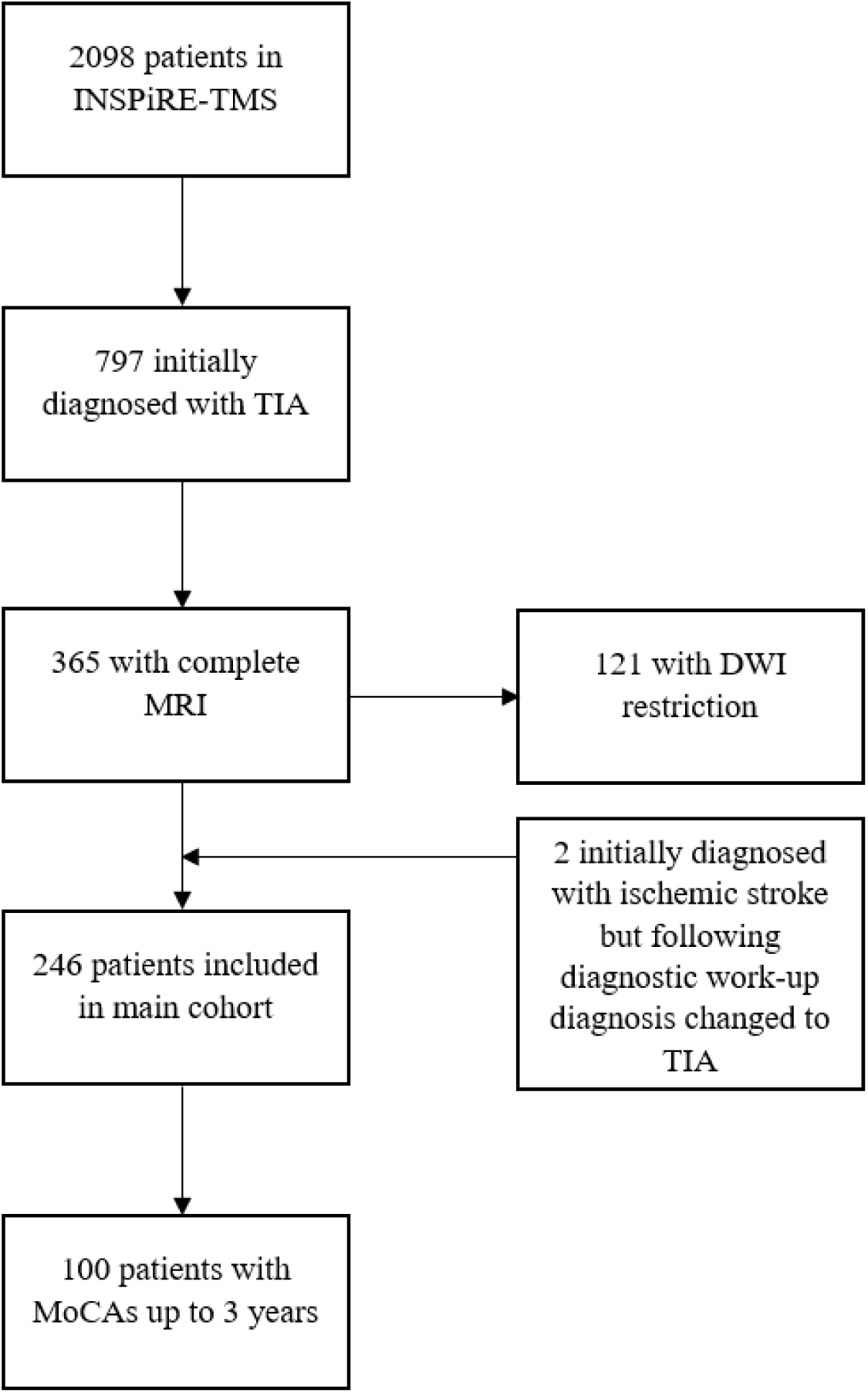
Patient selection flow chart. (INSPiRE-TMS = Intensified Secondary Prevention Intending a Reduction of Recurrent Events in TIA and Minor Stroke Patients, TIA = transient ischemic attack, MRI = magnetic resonance imaging, DWI = diffusion-weighted imaging, MoCA = Montreal Cognitive Assessment)

### Outcome Assessment

Cognitive status was assessed using the MoCA score; the score ranges from 0 to 30 in which higher scores represent better cognitive performance.^15^ It assesses the subdomains visuospatial abilities/ executive functions, naming, attention, abstract thinking, short-term memory and orientation to time and place.^16^ MoCA was assessed at baseline (during the acute hospital stay), and at annual follow-up outpatient visits up to 3 years post-stroke.^14^ All analyses involving assessment of cognitive decline were adjusted for age.

### Cerebral Small Vessel Disease Score on Baseline MRI

Imaging was performed locally with a 3 Tesla MRI scanner (Tim Trio; Siemens AG, Erlangen, Germany). To assess acute diffusion restriction, as well as the CSVD-score, we used axial diffusion-weighted imaging (DWI; scanning parameters: TE = 93.1 ms, TR = 7600 ms, FOV = 230 mm, matrix = 192 × 192, 2.5 mm section thickness with no intersection gap), T1- and T2*-weighted imaging (scanning parameters: TE = 20.0 ms, TR = 620.0 ms, FOV = 220 mm, matrix = 256 × 192, 5.0 mm section thickness with 0.5 mm intersection gap) and fluid attenuated inversion recovery (FLAIR) sequences (scanning parameters: TE = 100 ms, TR = 8000 ms, FOV = 220 mm, matrix = 256 × 256, 5.0 mm section thickness with 0.5 mm intersection gap).

The MRIs were initially assessed independently by two board certified radiologists. CSVD features were rated independently by two raters (P.R. and H.F.A.) blinded to cognitive outcomes. All ratings were supervised by a physician and imaging expert (A.K.).

The CSVD-score is a score ranging from 0 to 4 where points are allocated for the presence of CMBs, lacunes and PVS as well as relevant white matter hyperintensities (WMH).^17^ Cerebral microbleeds were defined as small areas of signal void with associated blooming on T2* sequences.^18^ The CMB distribution was categorized according to the Microbleed Anatomical Rating Scale.^19^ The WMH burden was assessed in FLAIR sequences with the Age-Related White Matter Changes (ARWMC) score by Wahlund et al.^20^ The cutoff was set at 10 with ARWMC <10 as non-relevant and MRIs with ARWMC ≥10 were given a point in the CSVD-score for significant WMH.^21^ The presence of lacunes, as well as the presence of PVS within the basal ganglia or centrum semiovale were assessed on FLAIR sequences using the STRIVE-2 criteria.^18^ In cases in which T1 or T2* were not available, B0-DWI was used instead.

### Statistical Analysis

To assess factors associated with cognitive decline up to 3 years following the acute cerebrovascular event (TIA), we performed a linear mixed model in which the dependent variable was the MoCA score (continuous) and random intercepts for individuals. In the first linear mixed model, intervention group, age, sex, time-point of assessment (baseline, 1-year, 2-year, 3-year) and total CSVD-score (range 0-4) were included as fixed effects. Models with and without interaction between time point and total CSVD score were compared using Akaike Information Criterion (AIC) and Bayesian Information Criterion (BIC); the best model was selected based on the lowest AIC and BIC values. In this study, the best model was identified as the model in which no interaction effect between time point and CSVD score was assessed.

In a linear mixed model for MoCA (continuous), the CSVD total score was replaced by individual CSVD component scores, namely CMB count (continuous), presence of enlarged PVS (binary), presence of lacunes (binary), and total WMH burden defined by ARWMH (continuous). To assess potential collinearity among predictors, we ran separate mixed models for individual predictors and compared the results with those from the full model. The consistency of the parameter estimates across these analyses indicated that collinearity was not an issue in our model. For all mixed models, the subjects were included as a random effect (random intercept). Furthermore, we performed a mixed model analysis for new onset MCI (mild cognitive impairment, defined as MoCA<26) including the following covariates: CSVD-score, age, timepoint, sex and intervention group as fixed effects.^16^

In order to assess whether total CSVD-score leads to differential decline across cognitive sub-domains, we z-normalized the scores within each of the six cognitive sub-domains including *visuospatial/ executive, naming, attention, abstraction, memory* and *orientation*.^16^ Subsequently, individual linear mixed models (random intercept models with random intercepts for individuals) were run on each normalised subcategory score used as the dependant variable; fixed effects included intervention group, age, sex, time-point of assessment (baseline, 1-year, 2-year, 3-year) and total CSVD-score; subjects were included as a random effect (random intercepts).

A two-sided significance level of α = 0.05 was used. Due to the exploratory nature of the current analysis, no adjustment was made for multiple testing therefore p-values should be interpreted with caution. Interpretation of findings is based on effect estimates and corresponding 95% confidence intervals (CI). All statistical analyses were performed using Stata (StataCorp Version 17.0).

## Results

### Patient Cohort Description

Of the 246 patients included in the current study, mean age was 69.4 years (± standard deviation [SD] 10.4) and 103 patients (41.9 %) were female. Median ABCD2 score at baseline was 4 (interquartile range [IQR] 3-5), median National Institutes of Health Stroke Scale (NIHSS) at the time of admission to the stroke unit was 0 (IQR 0-1) and baseline median mRS was 1 (IQR 1-1). No patients had a MoCA of <26 at baseline. The subgroup of 100 patients with available MoCAs up to 3 years follow-up had similar patient demographics in terms of age (mean age 67.1 ±SD 10.5) and distribution of sex (43 % female). Severity of initial symptoms in terms of baseline NIHSS and mRS as well as cardiovascular risk factor distribution was similar to the entire patient’s cohort. Detailed description of patient characteristics of the entire analyzed cohort and subgroup are described in **Table 1**.

**Table 1:** Patient cohort characteristics.

|  | Total patient cohort (n=246) | MoCA subgroup (n=100) | Non-MoCA subgroup (n=146) |
| --- | --- | --- | --- |
| **Demographics** |  |  |  |
| Age, mean (±SD) | 69.4 (10.4) | 67.1 (10.5) | 70.8 (10.1) |
| Sex, female, n (%) | 103 (41.9) | 43 (43.0) | 61 (41.8) |
| Education level ≥ 10 years, n (%) | 174 (72.8) | 72 (74.2) | 103 (72.5) |
| **Baseline clinical characteristics** |  |  |  |
| ABCD2 Score at admission, Median (IQR) | 4 (3-5) | 4 (3-4.5) | 4 (3-5) |
| NIHSS at admission, Median (IQR) | 0 (0-1) | 0 (0-1) | 0 (0-1) |
| mRS at admission, Median (IQR) | 1 (1-1) | 1 (1-1) | 1 (1-1) |
| TOAST criteria, n (%) |  |  |  |
| Large-artery atherosclerosis | 15 (6.1) | 3 (3.0) | 11 (7.6) |
| Cardioembolic stroke | 39 (15.9) | 15 (15.0) | 24 (16.6) |
| Small-vessel occlusion | 10 (4.1) | 3 (3.0) | 7 (4.8) |
| Other determined etiology | 1 (0.4) | 1 (1.0) | 0 (0) |
| Undetermined etiology | 180 (73.5) | 78 (78.0) | 103 (71.0) |
| **Cardiovascular risk factors** |  |  |  |
| Hypertension, n (%) | 211 (93.7) | 88 (91.7) | 123 (95.4) |
| Diabetes, n (%) | 57 (25.3) | 20 (20.8) | 37 (28.7) |
| Hypercholesterinemia, n (%) | 203 (90.6) | 87 (90.6) | 116 (90.6) |
| Atrial fibrillation, n (%) | 50 (22.4) | 19 (20.2) | 31 (24.0) |
| Current smoking, n (%) | 25 (10.2) | 7 (7) | 18 (12.3) |
| MoCA = Montreal Cognitive Assessment, SD = standard deviation, IQR = interquartile range, NIHSS = National Institutes of Health Stroke Scale, mRS = modified Rankin Scale, TOAST = Trial of Org 10172 in acute stroke treatment |  |  |  |

### Prevalence of Cerebral Small Vessel Disease

In the entire cohort (n=246), 58.1 % had a CSVD-score of ≥1; 28.5 % of the patients had a score of 1, 19.5 % a score of 2, 7.7% a score of 3 and 2.9% a score of 4. The most prevalent CSVD marker was the presence of lacunes (36.6 %), followed by enlarged PVS (28.1 %), WMH (19.5 %) and CMBs (17.9 %).

The CMB count ranged from 0 to 15 and the most common location was lobar (52.3 %), followed by mixed locations with CMB scattered across lobar, deep gray matter and infratentorial regions (36.4 %). The median ARWMC score was 4 (IQR 0-8) and 19.5 % of the participants had a high enough score (ARWMC ≥ 10) to be relevant for the CSVD-score. Full CSVD imaging characteristics of the entire analyzed cohort and subgroup are described in **Table 2**.

**Table 2:** Breakdown of CSVD marker distribution and CSVD severity in INSPiRE-TMS TIA patient cohort.

|  | Total patient cohort (n=246) | MoCA subgroup (n=100) | Non-MoCA subgroup (n=146) |
| --- | --- | --- | --- |
| **Cerebral microbleeds** |  |  |  |
| Present, n (%) | 44 (17.9) | 17 (17.0) | 27 (19.2) |
| Count: |  |  |  |
| 1 | 15 (6.2) | 6 (6.0) | 9 (6.2) |
| 2-4 | 18 (7.3) | 6 (6.0) | 12 (8.2) |
| > 4 | 11 (4.5) | 5 (5.0) | 6 (4.8) |
| **Location:** |  |  |  |
| Deep | 0 (0) | 0 (0) | 0 (0) |
| Infratentorial | 5 (11.4) | 3 (17.7) | 2 (7.0) |
| Lobar | 23 (52.3) | 5 (29.4) | 18 (64.3) |
| Mixed | 16 (36.4) | 9 (52.9) | 7 (28.6) |
| **White matter hyperintensities** |  |  |  |
| ARWMC score, Median (IQR) | 4 (0-8) | 4 (0-8) | 5 (4-8) |
| Relevant for CSVD-score, n (%) | 48 (19.5) | 20 (20.0) | 28 (19.2) |
| **Lacunes** |  |  |  |
| Present, n (%) | 90 (36.6) | 33 (33.0) | 57 (39.0) |
| **Perivascular spaces** |  |  |  |
| Present, n (%) | 69 (28.1) | 21 (21.0) | 47 (32.2) |
| **Cerebral small vessel disease** |  |  |  |
| CSVD-score, Median (IQR) | 1 (0-2) | 0 (0-2) | 1 (0-2) |
| Total CSVD-score, n (%) |  |  |  |
| 0 | 102 (41.5) | 52 (52.0) | 50 (34.3) |
| 1 | 70 (28.5) | 21 (21.0) | 49 (33.6) |
| 2 | 48 (19.5) | 15 (15.0) | 33 (22.6) |
| 3 | 19 (7.7) | 8 (8.0) | 11 (7.5) |
| 4 | 7 (2.9) | 4 (4.0) | 3 (2.1) |
| CSVD = cerebral small vessel disease, IQR = interquartile range, INSPiRE-TMS = Intensified Secondary Prevention Intending a Reduction of Recurrent Events in TIA and Minor Stroke Patients, TIA = transient ischemic attack, ARWMC = Age-Related White Matter Changes |  |  |  |

### CSVD and Cognitive Trajectory

Out of 64 patients with a 3-year follow-up, 42 patients (65.6%) had a mild cognitive impairment, defined by MoCA < 26 at baseline. In the linear mixed model analysis for MoCA (continuous) assessed up to 3 years following TIA, total baseline CSVD-score (range 0-4) and age were independently associated with overall MoCA score with an adjusted β of -0.53 (95% CI [confidence interval] -0.97 – -0.09, p = 0.018), and -0.08 (95% CI -0.13 – -0.03, p=0.001), respectively (**Table 3**). Timepoint, sex and randomization group were not significantly associated with cognitive outcomes in this model (**Table 3**). In a mixed model analysis for new onset MCI (MoCA <26), CSVD-score had an adjusted β of -0.37 (95% CI -0.79 – 0.03, p = 0.071; **Supplementary Table 2**).

**Table 3:**
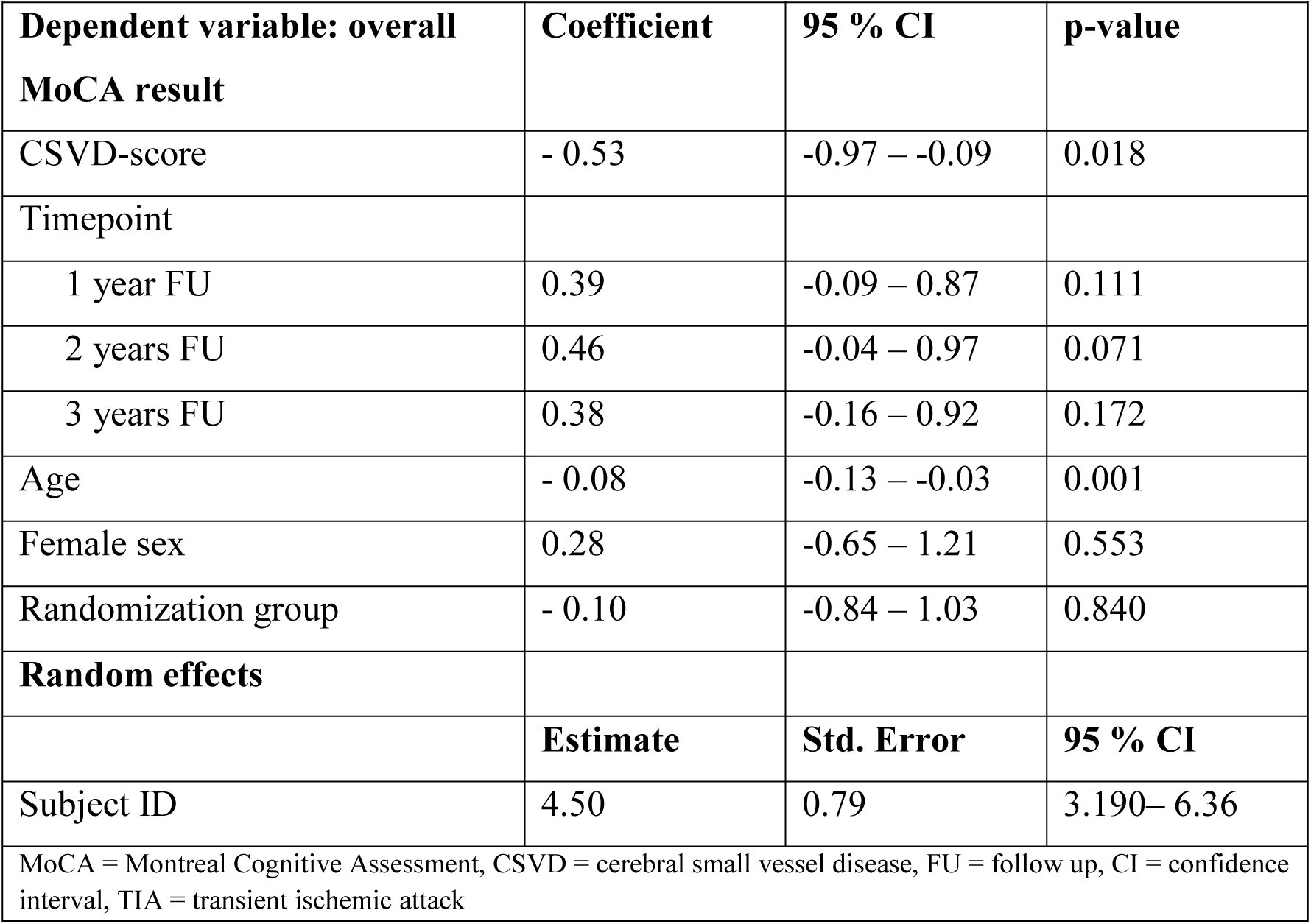
Linear mixed model for MoCA (continuous) assessed up to 3 years following TIA including intervention group, age, sex, time-point of assessment (baseline as reference) and total CSVD-score (0-4) as fixed effects (n_patients_ = 100, n_observertions_ = 329).

| **Dependent variable: overall**<br>**MoCA result** | **Coefficient** | **95 % CI** | **p-value** |
| --- | --- | --- | --- |
| CSVD-score | - 0.53 | -0.97 – -0.09 | 0.018 |
| Timepoint |  |  |  |
| 1 year FU | 0.39 | -0.09 – 0.87 | 0.111 |
| 2 years FU | 0.46 | -0.04 – 0.97 | 0.071 |
| 3 years FU | 0.38 | -0.16 – 0.92 | 0.172 |
| Age | - 0.08 | -0.13 – -0.03 | 0.001 |
| Female sex | 0.28 | -0.65 – 1.21 | 0.553 |
| Randomization group | - 0.10 | -0.84 – 1.03 | 0.840 |
| **Random effects** |  |  |  |
|  | **Estimate** | **Std. Error** | **95 % CI** |
| Subject ID | 4.50 | 0.79 | 3.190– 6.36 |
| MoCA = Montreal Cognitive Assessment, CSVD = cerebral small vessel disease, FU = follow up, CI = confidence interval, TIA = transient ischemic attack |  |  |  |

In an additional linear mixed model analysis for MoCA (continuous) assessed up to 3 years after TIA, CSVD sub-components were included as individual fixed effects (**Table 4**). A significant negative association was found between CMB count and MoCA scores, with an adjusted β of -0.42 (95% CI -0.63 – -0.21, p < 0.001; **Table 4**). In this model, age was also negatively associated with MoCA with an adjusted β of -0.09 (95% CI -0.13 – -0.04, p = 0.001). The other components of the CSVD-score were not significantly associated with the total MoCA score (**Table 4**).

**Table 4:** Linear mixed model for MoCA (continuous) assessed up to 3 years post TIA including intervention group, age, sex, time-point of assessment (baseline as reference) and individual CSVD component scores (CMB count, ARWMC score, presence of lacunes and PVS) as fixed effects (n_patients_ = 100, n_observertions_ =329).

| **Dependent variable: overall**<br>**MoCA result** | **Coefficient** | **95 % CI** | **p-value** |
| --- | --- | --- | --- |
| CMB count (continuous) | - 0.42 | -0.63 – -0.21 | < 0.001 |
| ARWMC score (continuous) | 0.00 | -0.12 – 0.12 | 0.996 |
| Lacunes present (binary) | - 0.59 | -1.75 – 0.57 | 0.320 |
| PVS present (binary) | 0.50 | -0.63 – 1.62 | 0.385 |
| Timepoint |  |  |  |
| 1 year FU | 0.39 | -0.09 – 0.87 | 0.111 |
| 2 years FU | 0.48 | -0.02 – 0.98 | 0.060 |
| 3 years FU | 0.38 | -0.16 – 0.91 | 0.170 |
| Age | - 0.09 | -0.13 - -0.04 | 0.001 |
| Female sex | 0.53 | -0.37 – 1.42 | 0.248 |
| Randomization group | - 0.08 | -0.80 – 0.95 | 0.867 |
| **Random effects** |  |  |  |
|  | **Estimate** | **Std. Error** | **95 % CI** |
| Subject ID | 3.83 | 0.69 | 2.69 – 5.44 |
| MoCA = Montreal Cognitive Assessment, CMB = cerebral microbleed, ARWMC = Age-Related White Matter Changes, PVS = perivascular spaces, FU = follow up, CI = confidence interval, TIA = transient ischemic attack, CSVD = cerebral small vessel disease |  |  |  |

### Impact of CSVD-Score on Cognitive Sub-domains

For all sub-domains of cognition except for orientation, higher CSVD-scores were associated with worse domain-specific performance (**Figure 2**), however the association with the total CSVD-score was strongest for the *memory* domain (β = -0.18, 95% CI -0.32 – -0.04, p = 0.014). For the *visuospatial/ executive* domain, total CSVD-score had an adjusted β of -0.12 (95% CI -0.26 – 0.02, p =0.093); for *naming*, an adjusted β of -0.09 (95% CI -0.19 – 0.01, p = 0.081); for *attention*, an adjusted β of -0.06 (95% CI -0.21 – 0.09, p = 0.421) and for *abstract thinking*, an adjusted β of -0.03 (95% CI -0.15 – 0.09, p = 0.630). For *orientation*, the CSVD-score had an adjusted β of 0.04 (95% CI -0.09 – 0.18, p = 0.541). Depending on which subdomain, confounding factors such as age and sex had differential effect sizes; the results of each specific mixed model can be found in **Supplementary Table 3**.

**Figure 2:**
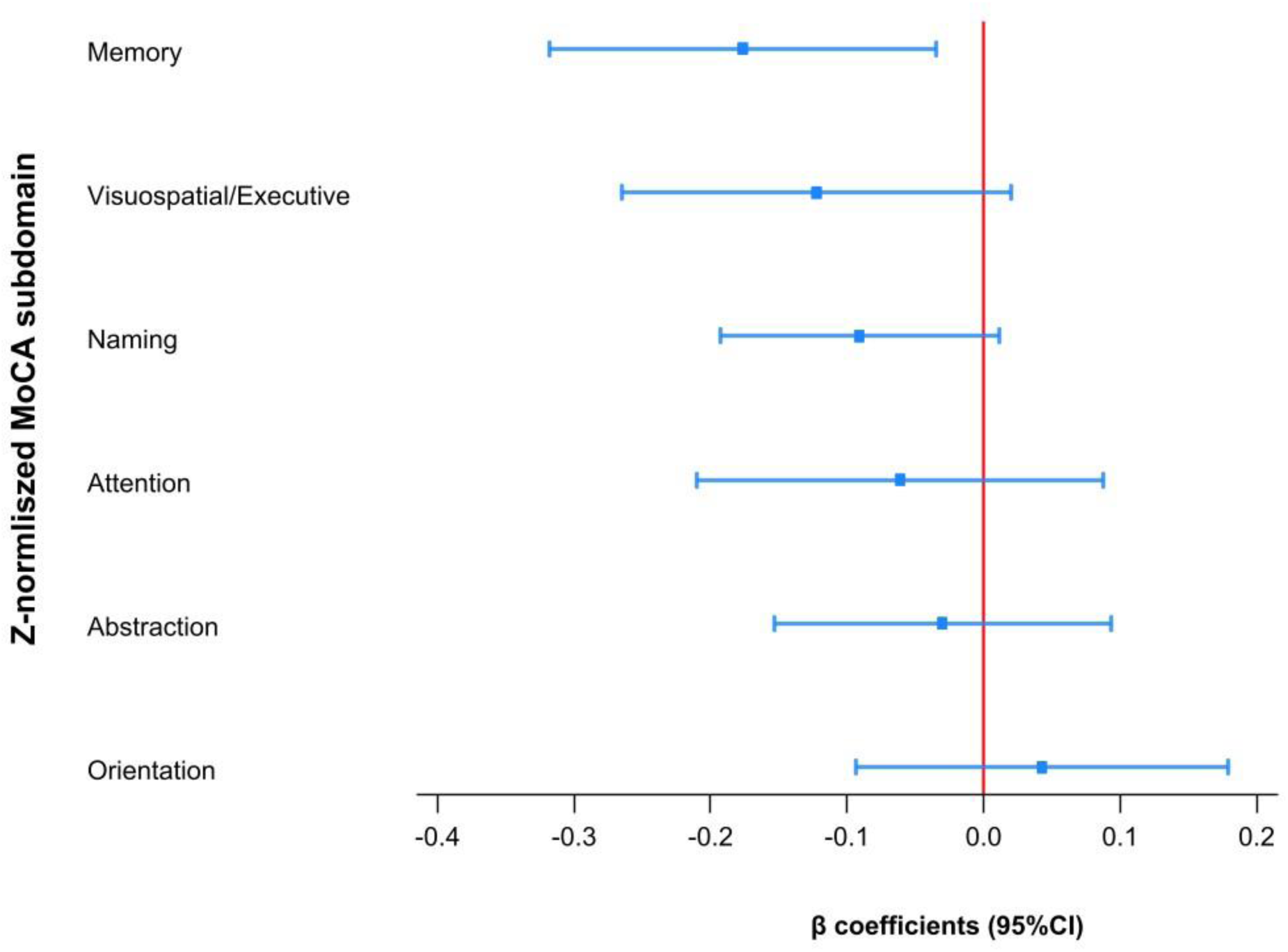
Coefficient plot of impact of CSVD-score (0-4) on each z-normalized MoCA subdomain assessed up to 3 years post TIA (n_patients_ = 100, n_observertions_ =329); β coefficients and 95% confidence intervals of linear mixed model (subcategory score as the dependant variable; fixed effects included intervention group, age, sex, time-point of assessment and total CSVD-score; subjects were included as a random effect) depicted. A negative β indicates that the more CSVD severity increases (i.e., higher CSVD-score), the more cognitive performance in the MoCA subdomain worsens (i.e., lower z-normalized score), whereas a positive β suggest that higher CSVD severity is associated with better cognitive performance. (CSVD = cerebral small vessel disease, MoCA = Montreal Cognitive Assessment, TIA = transient ischemic attack, CI = confidence interval)

## Discussion

In this study, we found that not only was CSVD present on MRI in nearly 60% of TIA patients, but that the severity of CSVD - as assessed via the CSVD-score - was independently associated with worse cognitive outcomes up to three years after the acute ischemic event, in addition to older age. Although CSVD severity was associated with poorer performance in the specific cognitive domain of *memory*, higher CSVD-scores were negatively associated with nearly all cognitive subdomains except *orientation*.

This study included 246 moderate-to-high-risk TIA patients (ABCD2 score ≥ 3) from the INSPiRE-TMS trial, with a mean age of 69 years. In terms of cerebrovascular risk profiles and stroke etiology, the study population is representative of a cardiovascular high-risk population.^22,23^ In the subgroup analysis of 100 TIA patients with available longitudinal MoCA data, patient demographics and clinical characteristics were similar to the overall cohort (**Table 1**).

Nearly 60% of the included TIA patients had a CSVD-score of ≥1, indicating the presence of at least one imaging biomarker of CSVD and incipient cerebrovascular disease (**Table 2**).^17^ All features of the CSVD-score were higher in older aged patients compared to younger patients (**Supplementary Table 1**). These findings align with existing literature that highlights the high prevalence of CSVD markers in populations with vascular risk factors and advanced age.^24,25^ Among the individual MRI features included in the total CSVD-score, lacunes were the most prevalent; studies have shown that the presence of lacunes not only increases stroke risk but may also be associated with increased cognitive impairment.^26^ Prominent PVS were the second most frequent MRI feature. Interestingly, previous studies have found that enlarged PVS may be an indicator of microvascular dysfunction and may be linked to cognitive decline.^27,28^ A high WMH burden (ARWMC ≥10) was present in a fifth of patients; WMH are a known risk factor for recurrent vascular events and cognitive decline in stroke populations.^29,30^ CMBs were visible in 18% of TIA patients and were predominantly located in lobar regions; CMBs have also been linked to increased risk of cognitive decline in ageing individuals and stroke patients.^31,32^

In a linear mixed model analysis, total CSVD-score was interpedently associated with worse cognitive outcomes up to 3 years following the ischemic event (adjusted β of -0.53, 95% CI - 0.97 – -0.09, p = 0.018; **Table 3**). Age was significantly associated with worse cognitive outcomes in this patient population (**Table 3**). These findings stand in line with the literature.^10^ Interestingly, CMB count was identified as the strongest component of the CSVD-score in terms of association with cognitive outcomes compared to the other CSVD biomarkers (adjusted β of -0.42, 95% CI -0.63 – -0.21, p < 0.001; **Table 4**). Although previous studies have found CMBs to be associated with cognitive dysfunction in mixed stroke and TIA populations^33–36^, most studies consider high WMH burden to be the strongest predictor for cognitive decline.^21,37,38^ Although white matter burden in our study population was moderate to high (median ARWMC 4 [IQR 0-8], 20% with high WMH burden ARWMC≥10), WMH burden alone was not associated with cognitive decline in this population of high-risk TIA patients. These findings – if validated in future independent studies - suggest that we may need to pay particular attention to the presence of CMBs in TIA patients, as these may identify a particularly high-risk group for long-term cognitive dysfunction in patients after TIA.

Severity of CSVD (total CSVD-score) was negatively associated with nearly all cognitive subdomains except for *orientation*. In this exploratory analysis, we observed that total CSVD-score was significantly associated with worse *memory* function up to 3 years (**Figure 2**). This aligns with the hypothesis that vascular contributions to cognitive impairment disproportionately affect areas related to memory and executive functioning; possibly due to their reliance on the integrity of distributed neural networks vulnerable to ischemic damage.^5,39^ There was a moderate negative association between a higher CSVD-score and *visuospatial/ executive functions, naming, attention* and *abstract thinking*, suggesting these domains may also be compromised with higher vascular burden (**Figure 2**)^40^. In contrast, no substantial association was observed for the cognitive subdomain *orientation* likely due to its relative resilience in early cognitive dysfunction and the ceiling effect of orientation tasks within MoCA, which may not be sensitive enough to detect subtle impairments. Ultimately, damage caused by progressive CSVD is likely to cause differential decline in cognitive subdomains depending on severity, and the presence/absence of component features (e.g., WMH burden vs. CMB count). Future larger, independent studies focusing exclusively on TIA patients are needed to validate these findings and further investigate subdomain-specific cognitive decline based on MR-biomarkers of CSVD.

There are several limitations of the current study that warrant discussion. First and foremost, the post-hoc and observational design of the study increases the risk of Type II error and limits the ability to establish causality between CSVD and cognitive outcomes. Furthermore, the rather small sample size reduces capability to detect subtle effects. There was also no adjustment made for multiple testing in this exploratory study. Additionally, there is a selection bias, as only patients who received an MRI were included in the current analysis, hereby automatically excluding all patients with contraindications for MRI; patients with MRI contraindications often have more comorbidities, including cardiac pre-existing conditions.^41^ In other words, generalizability of our findings may be limited. Furthermore, over 30% of the cohort had lacunes or chronic infarctions (location was not assessed), which could themselves contribute to cognitive impairments, complicating the interpretation of CSVD-specific effects.^39^ Additionally, the reliance on MoCA as the primary cognitive assessment tool, while a well-established and widely used screening tool for cognitive function, may have underestimated deficits in specific domains such as executive function and processing speed and might have a learning effect when used repeatedly.^42–44^ Larger prospective studies with more in-depth neuropsychological assessments of cognitive function are needed to validate these findings and address these limitations.

A major strength of this study is the inclusion of TIA patients without acute diffusion restriction on MRI exclusively, reducing the confounding acute lesion effect on cognition.^39^ In other words, this design allows for a more precise assessment of the association of CSVD alone on cognitive outcomes. Another strength of the study is the domain-specific cognitive analysis, combined with its longitudinal design with annual follow-ups up to three years following the index event. Lastly, this study uses patient data stemming from a large, multicenter randomized controlled trail with comprehensive, longitudinal long-term follow-up data.

In summary, biomarkers of CSVD are present in more than half of patients who experience a TIA. The most prevalent imaging biomarker of CSVD in TIA patients were lacunes, followed by PVS, WMH and CMBs. Cumulative CSVD-score assessed on baseline MRI was independently associated with worse cognitive performance up to 3 years following the acute ischemic event, alongside increased age. Interestingly, CMB burden was the strongest component of the CSVD-score in terms of association with worse cognitive outcomes in this high-risk TIA patient population. The severity of CSVD, as reflected by the cumulative score of imaging biomarkers, had the strongest negative effect on the *memory* sub-domain of cognition. We also observed a negative, moderate association for all other cognitive subdomains except for *orientation*, suggesting that CSVD severity likely has a differential impact on cognitive sub-domains. This study highlights the importance of patient individual risk stratification in patients with TIA and visible CSVD biomarkers on MRI. If validated in future, independent, larger studies, early CSVD assessment could be used as an important tool to initiate secondary prevention strategies to prevent cognitive decline in high-risk TIA populations.

## Acknowledgements

M.E. received funding from DFG under Germanýs Excellence Strategy – EXC-2049 – 390688087, Collaborative Research Center ReTune TRR 295-424778381, Clinical Research Group KFO 5023 BeCAUSE-Y, project 2 EN343/16-1. BMBF, DZNE, DZHK, DZPG, EU, Corona Foundation, and Fondation Leducq

## Author Contribution Statement

A.K. and P.R. jointly conceived the study. A.K., P.R., U.G. and P.G. designed and performed data analysis and interpreted the results. P.R. wrote the manuscript. P.R. pre-processed the MRI data. A.K., U.G. and P.G. gave conceptual and analytical advice. H.F.A. gathered neuroimaging data and helped with image pre-processing. A.K. performed language assessment and preprocessed the data. U.G. and P.G. helped with statistical analyses. HJA was the Principal Investigator of the INSPiRE-TMS study. All authors discussed the results and critically revised the manuscript.

## Data Availability Statement

Data supporting the results of this study can be provided upon request.

